# Impact of a Non-Mandatory Electronic Medical Record Protocol on Venous Thromboembolism Prophylaxis in Adult Hospitalized Patients: A Before-and-After Cohort Analysis of 177,856 Cases

**DOI:** 10.1101/2025.08.22.25334227

**Authors:** Marcelo Passos Teivelis, Bruno Jeronimo Ponte, Emanuelle Lima Macedo, Isabela Monforte de Toledo, Mateus de Lima Freitas, Adriano Jose Pereira, Maria Fernanda Casino Portugal, Valéria Pinheiro de Souza, João Carlos de Campos Guerra, Nelson Wolosker

## Abstract

**Background:** Venous thromboembolic events (VTE), which include deep vein thrombosis (DVT) and pulmonary embolism (PE) are common conditions within hospitalized patients, and is a well-known cause of in-hospital morbidity and mortality. The evidence on thromboprophylaxis benefits for reduce VTE in hospitalized patients is unequivocal. Despite that, evidence on electronic decision supports systems (e-DSS) and the real impact in VTE prevalence is still lacking.

**Methods:** A retrospective analysis comprising all admissions between 2017 to 2021, two years before, and two years after the implementation of the e-DSS in a quaternary hospital was carried out. Patients were divided in two groups: pre-e-DSS and post-e-DSS. VTE cases were defined as any occurrence of DVT in the lower or upper limbs, superficial phlebitis, or PE. A subgroup analysis was realized based on patients’ categories of ICD-10 from admission records, including Surgical, Orthopedic, Obstetrical and Clinical admissions. The prevalence of VTE in the pre-e-DSS and post-e-DSS was compared.

**Results:** A total of 177,851 medical records were included in the analysis. The pre-e-DSS group consisted of 104,943 admissions (59%), while the post-e-DSS 72,913 (41%). A total of 1,059 (0,60%) events of thrombosis were diagnosis in hospitalized patients. In the pre-e-DSS group 584 VTE were diagnosed, the post-e-DSS demonstrated 475. No statistical difference was demonstrated between both groups (p 0,011). The subgroup analysis considering patients from Surgical (p 0,524), Orthopedic (p 0,034) and Obstetrical (p 0,870) admissions demonstrated no statistical difference. The Clinical admissions demonstrated an increase in VTE in the group post-e-DSS (p<0,001).

**Conclusion:** The use of a non-mandatory electronic protocol for managing venous thromboembolism in patients’ electronic records is insufficient to reduce VTE incidence among hospitalized patients across all groups, including Surgical, Clinical, Orthopedic, and Obstetrical.

## INTRODUCTION

Venous thromboembolic events (VTE), which include deep venous thrombosis (DVT) and pulmonary embolism (PE), are common conditions within the population. (1,2) They are a well-known cause of in-hospital complications and have consistently been identified as the leading cause of preventable in-hospital mortality. (1,3)

Management of VTE primarily relies on the use of anticoagulants; (4) however, certain cases may require the implantation of a vena cava filter, percutaneous mechanical thrombectomy, or systemic or catheter-guided thrombolysis. (5–8) A population-based study by Stein et al. demonstrated that in a population of 612,000,000 hospital admissions in a 21-year period, the incidence of in-hospital VTE was 1,24%. (9) Given that even distal thrombosis in the lower limbs presents a significant risk of progressing to PE, early diagnosis and VTE prophylaxis are critical components of hospital medical practice. (1,10,11)

The evidence supporting the use of thromboprophylaxis for appropriate groups of at-risk patients is unequivocal, with multiple randomized clinical trials validating its safety, efficacy and cost-effectiveness. (3,12,13) Despite the consistent publication and updating of guidelines on the topic (12,14), the implementation of preventive measures varies widely among healthcare institutions, with notable variations even within the same services(15). Evidence indicates persistent shortcomings in the application of thromboprophylaxis in both European (16) and North American institutions over the past decade. (17–19)

In a 2018 meta-analysis, Kahn et al. found that prophylaxis rates improved when a warning tool was incorporated into the electronic medical record (EMR), particularly when it was used at the time of prescription (20). Electronic alerts appeared to be more effective; however, a specific analysis was not feasible due to a lack of trials. (20) Added to that, in 2024 a retrospective analysis carried out by Bahl et al. documented, in a large casuistry, that adherence to filling out protocols for VTE prophylaxis increased after it became mandatory. (21)

In our institution, with the objective of diminishing the risk of VTE in hospitalized patients, an electronic Decision Support System (e-DSS) has been operational since 2019, providing prophylaxis warnings and recommendations when a patient’s EMR is accessed. However, using the e-DSS is not mandatory, and it does not restrict access to other parts of the medical record.

To the best of our knowledge, only one North American study has analyzed the completion of electronic VTE protocols, linking them to the prevalence of VTE in hospitalized patients, using a substantial sample. (21)

Therefore, this study aimed to retrospectively assess the rates of VTE confirmed by radiological exams in a large population (177.856 patients) before and after the implementation of the e-DSS at a quaternary care hospital with nearly 700 operational beds in São Paulo, Brazil.

## METHODS

### Design

This was a retrospective analysis of patient records, including all admissions between 2017 and 2021 in a Brazilian hospital. The objective of this study was to assess whether there was a difference in VTE among inpatients after implantation of an electronic Decision Support System (e-DSS). This study was approved by the Institutional Ethics Committee under protocol number 45305121.5.0000.0071, and an informed consent form was waived for the patients included.

### Population

The study population consisted of all admissions recorded between 2017 and 2021, two years before and three years after the implementation of the e-DSS in an EMR. Patients were divided into two groups: the pre-e-DSS Group, which included those admitted before the implementation of the e-DSS, and the post-e-DSS Group, which comprised patients admitted after 2019, when the system became operational.

Exclusion criteria established were patients under 18 years of age, those admitted with a prior outpatient VTE diagnosis without hospitalization in the preceding 30 days, and individuals diagnosed with COVID-19 within 30 days before admission, during hospitalization, and 30 days after discharge. To identify patients with exclusion factors related to COVID-19, confirmatory laboratory tests and ICD-10 codes related to hospitalization were analyzed. Any patient presenting with these exclusion criteria was removed from the sample.

### VTE Definition and Stratification

VTE cases were defined as any occurrence of DVT in the lower or upper limbs, superficial phlebitis, or PE. Positive cases were automatically cross-referenced with the complete dataset using medical record numbers.

Data from the radiology department included venous Doppler ultrasound scans of the lower and upper limbs, as well as pelvis and chest CT reports. These reports were screened for terms such as *thrombosis, thrombophlebitis*, and *thromboembolism*.

Two independent authors reviewed all flagged reports to confirm the presence of VTE. If a patient had multiple positive imaging tests, only the earliest confirmed diagnosis was considered, and subsequent results were excluded.

### e-DSS application

The e-DSS was introduced in 2019 as an add-on to the existing electronic medical records system, Cerner Millennium^®^ (Cerner Corporation, North Kansas City, USA), in use since 2016. This tool serves as a clinical decision support mechanism, automatically applied to all hospitalized patients to calculate VTE risk scores and recommend appropriate prophylaxis, following the 2016 CHEST AT9 guidelines update. (22) The attending physician could accept or refuse to fill in the e-DSS. Consequently, they could accept or deny the recommendation provided by the e-DSS regarding VTE prophylaxis, providing a justification if they chose not to accept it. This study did not assess adherence to the system’s recommendations.

### Subgroup Analysis

The patients were classified into four categories based on their International Classification of Diseases (ICD-10) codes: clinical, surgical, orthopedic, and obstetric. Two of the authors were responsible for classifying each ICD into its respective category. The ICD codes assigned to each group are provided in the Supplementary Material (Supplement 1).

We first analyzed the prevalence of VTE in both groups, followed by an analysis of the subgroups categorized by ICD.

### Data and Statistical Analysis

Ordinal variables were presented as total and proportional frequencies. All data were considered to be normally distributed; therefore, we used averages and standard deviations as measures of dispersion.

Because the Pre-e-DSS Group and Post-e-DSS Group were considered dependent, the chi-square and Fisher’s exact test were used for comparison of quantitative features. All statistical analyses were conducted using the SPSS software for Windows, version 22.0 (IBM Corp., Armonk, NY). The p-value < 0.001 was established to conclude statistical significance.

## RESULTS

From 2017 to 2021, a total of 230,557 hospital admissions were analyzed. Out of these, 177,856 were included in the sample that was analyzed. (Figure 1) Specifically, 104,943 (59%) were classified as the pre-e-DSS group, while 72,913 (41%) admissions were classified as the post e-DSS group.

**Figure 1.**
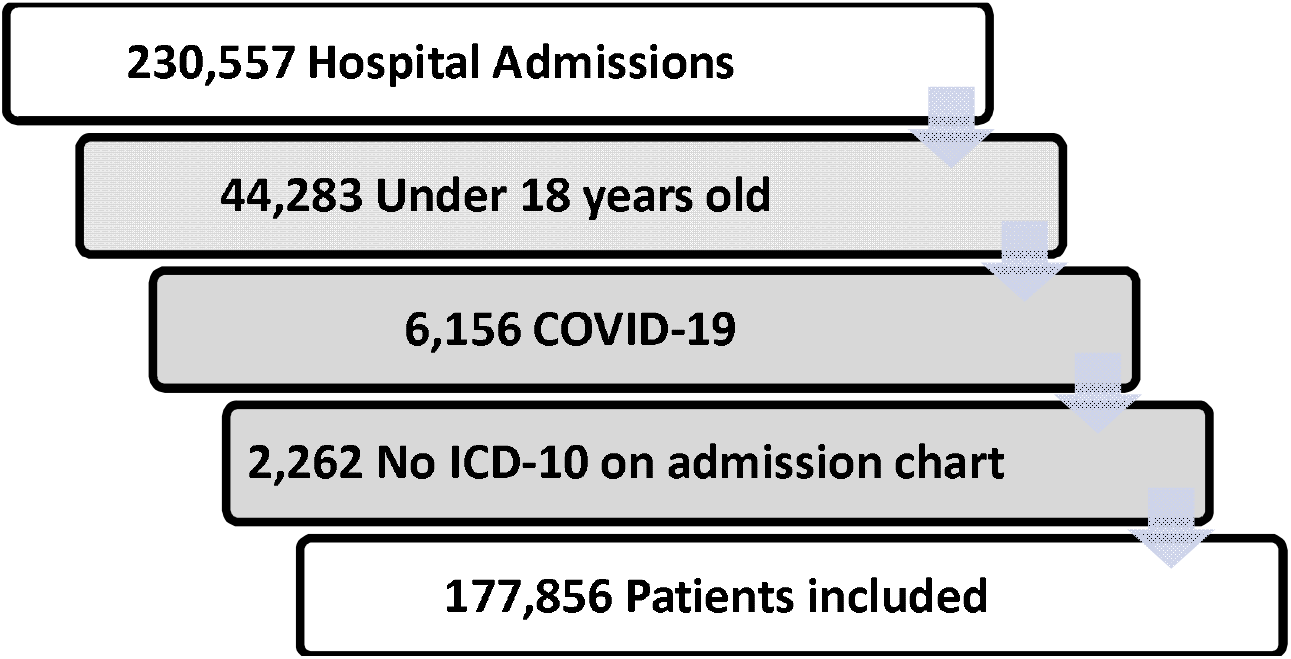
Patient enrolment flowchart

During the analyzed period, 4,823 radiological exams documented a VTE event. Out of these exams, the prevalence of VTE among inpatients was found to be 1,059 cases, which represents 0.60% of the total. Table 1 provides a detailed distribution of thromboembolic events in the pre- and post-implementation DSS populations.

**Table 1.**
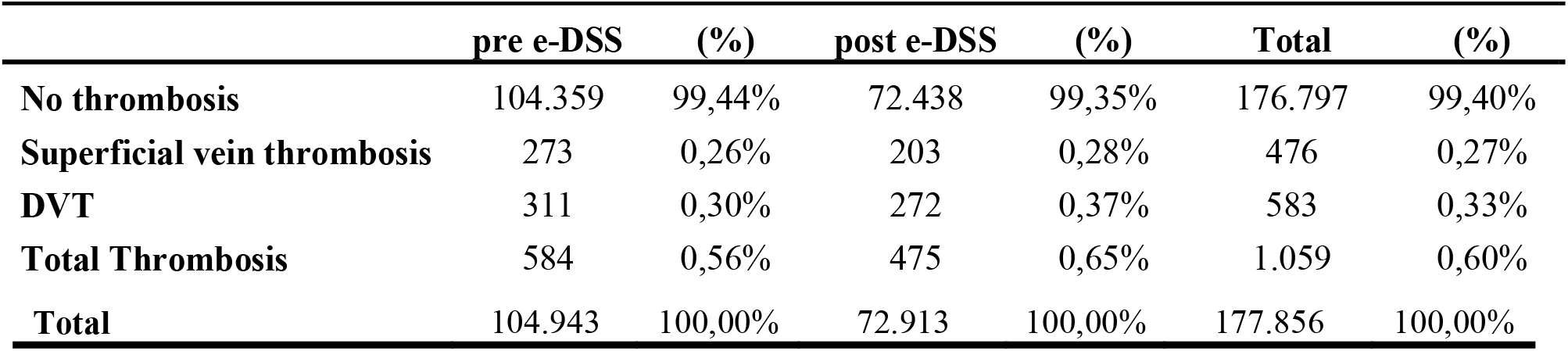
Distribution of thromboembolic events in pre- and post e-DSS implementation. DSS: Decision Support System.

|  | pre e-DSS | (%) | post e-DSS | (%) | Total | (%) |
| --- | --- | --- | --- | --- | --- | --- |
| No thrombosis | 104.359 | 99,44% | 72.438 | 99,35% | 176.797 | 99,40% |
| Superficial vein thrombosis | 273 | 0,26% | 203 | 0,28% | 476 | 0,27% |
| DVT | 311 | 0,30% | 272 | 0,37% | 583 | 0,33% |
| Total Thrombosis | 584 | 0,56% | 475 | 0,65% | 1.059 | 0,60% |
| **Total** | 104.943 | 100,00% | 72.913 | 100,00% | 177.856 | 100,00% |

The analysis of subgroups categorized by ICD is presented in Table 2. In the clinical admissions group, there was a slight increase in the rate of thrombotic events over the years (p<0.001). In contrast, the other groups did not show any significant differences following the implementation of the e-DSS.

**Table 2.** Composition of pre- and post e-DSS superficial and deep vein thrombosis events in the ICD-10 subgroups and total. DSS: Decision Support System.

|  | Pre- e-DSS |  | Post e-DSS |  | Chi2 | Odds Ratio | p-value |
| --- | --- | --- | --- | --- | --- | --- | --- |
|  | No Thrombosis | Thrombosis | No Thrombosis | Thrombosis |  |  |  |
| **Surgical** | 51,761 (99,57%) | 222 (0,43%) | 36,236 (99,60%) | 145 (0,40%) | 0.354 | 0.933 | 0.524 |
| **Clinical** | 27,526 (98.96%) | 289 (1.04%) | 17,997 (98.59%) | 258 (1.41%) | 12.844 | 1.365 | <0.001 |
| **Orthopedics** | 12,709 (99.61%) | 50 (0.39%) | 9,468 (99.40%) | 57 (0.60%) | 4.446 | 1.53 | 0.034 |
| **Obstetrical** | 12,363 (99.81%) | 23 (0.19%) | 8,737 (99,83%) | 15 (0,17%) | 0.006 | 0.923 | 0.870 |
| **Total** | 104,359 (99.44%) | 584 (0.56%) | 72,438 (99.34%) | 475 (0.66%) | 6.396 | 1.171 | 0.011 |

Overall, the global analysis, which included all subgroups, indicated that there were no significant differences in the prevalence of VTE after the implementation of the e-DSS.

## DISCUSSION

Quaternary hospitals, characterized by their high complexity of care, receive patients with a wide variety of clinical conditions. This study analyzed all hospital admissions in a single hospital over a 5-year period, excluding COVID-19 cases and patients under the age of 18. The final sample comprised 177,856 admissions. All records were kept using a standardized EMR (Cerner Millenium^®^), guaranteeing data consistency and reliability. The patients were divided into 2 groups for comparative analysis: before and after the implementation of the e-DSS.

Of the total number of admissions, we identified 1,059 cases of VTE, corresponding to a prevalence of 0.60%, which is lower compared to the study conducted by Stein et al. where an incidence of 1.24% of VTE was documented. (9) This low prevalence of VTE clearly reflects the effectiveness of the preventive measures adopted at our institution, an indication of high-quality care performance. Despite these results, in line with our culture of continuous improvement, we have implemented a new clinical alarm system (e-DSS) seeking to minimize the prevalence of in-hospital VTE.

To improve adherence to prophylactic measures, VTE consensus guidelines have encouraged healthcare institutions to standardize educational programs for clinical staff, focusing on risk factors and prevention strategies. (3) Additionally, while e-DSS have been tested as an alternative to implementing VTE prophylaxis protocols, the scientific evidence regarding their effectiveness in clinical practice is inconsistent. Despite strong evidence that thromboprophylaxis is safe, effective and cost-effective in reducing VTE events, the impact of these records remains debatable. (20,23)

Randomized studies, such as the trial by Kucher et al., have shown a 41% reduction in the risk of VTE within 90 days after implementing voluntary electronic alert systems. These systems were also linked to an increase in both mechanical and pharmacological prophylaxis use. (24) Similarly, studies by Dexter et al. and Beeler et al. reported improved prophylaxis prescription rates following the introduction of voluntary electronic protocols that were triggered during patient transfers between hospital departments. (25,26) More recently, Baugh et al. observed an absolute increase of 3.8% in the use of pharmacological prophylaxis in emergency observation units following the adoption of a clinical decision support tool. (27)

On the other hand, some studies have reported less consistent results concerning the impact of increased prescription rates on statistically significant reductions in VTE events. For instance, the study by Roy et al. found no decrease in thrombotic events, despite a rise in the overall use of antithrombotic therapy, which did not necessarily correspond to appropriate prophylaxis measures. (28)

Our analysis demonstrated that the implementation of e-DSS did not alter the prevalence of in-hospital VTE. This discrepancy suggests that the mere presence of an electronic protocol does not guarantee the achievement of the intended clinical outcomes, specifically, a reduction in VTE. However, when performing a subgroup analysis, we found a slight increase in the prevalence of VTE over the years among hospitalizations for clinical conditions. Several factors may be associated with this finding, including an increase in the hospitalization of patients at higher risk for VTE, such as the elderly and those with cancer, as well as a decrease in the prescription of VTE prophylaxis within this subgroup.

Orthopaedical surgery without prophylaxis has a high risk of postoperative thrombosis, estimated between 40% and 70%, while implementing preventive measures greatly reduces substantively this risk, depending on the procedure.(29) Among the subgroups in our analysis, most VTE events occurred in the clinical category, surpassing even the surgical groups, which is surprising since VTE risk is usually linked mainly to surgery.(3,16,28) Although surgery remains a significant thromboembolic risk, increased attention to monitoring clinical cases can be just as, if not more, effective in lowering VTE episodes in hospitals. Therefore, reinforcing prophylaxis strategies in medical wards is essential to optimize VTE prevention throughout all levels of hospitalization.

The effectiveness of electronic protocols appears to be intrinsically linked to the level of adherence from attending physicians. (30) In the study by Lecumberri et al., the electronic system prompted physicians to complete a questionnaire regarding the reason for hospitalization and any history of VTE, which may have led to a more proactive and appropriate prescription of prophylaxis. (31)

On the other hand, studies that did not demonstrate a statistically significant improvement in appropriate prophylaxis rates, such as the one by Spirk et al., highlighted that, although the system prompted physicians to complete a risk assessment, it allowed the task to be postponed up to three times. As a result, many alerts were ignored, and appropriate prophylaxis rates did not improve. (32) Similarly, Nendaz et al. reported low utilization of the electronic risk assessment tool when its use was optional. This led the authors to conclude that more coercive or mandatory systems may be more effective in promoting adherence. (33) Added to that, the retrospective study published in 2024 by Bahl et al. reinforces this information, since it was documented that changing the VTE protocol to mandatory increased the rate of adherence by clinical staff physicians.(21)

At our institution, completion of the e-DSS is not mandatory, which may cause delays in its usage or prevent attending physicians from properly following the protocol.

Although studies have demonstrated improved adherence to VTE prophylaxis among hospitalized patients (31), the simple display of an optional reminder within electronic medical records may be insufficient to enhance protocol adherence and, consequently, reduce the prevalence of VTE in the in-hospital population. Therefore, the adoption of educational measures for clinical staff physicians, as well as the consideration of mandatory VTE risk assessment protocols, may be useful tools for increasing adherence to the VTE prophylaxis, reducing the thromboembolic events in hospitalized patients.

Educational initiatives and awareness campaigns on thromboembolic risks, together with the implementation of more mandatory or structured protocols for VTE, may be effective measures for reducing the prevalence of VTE events in hospitalized patients.

## LIMITATIONS

Given the large sample size, analyses were not performed on an individual basis, and subgroup categorization was based on the primary ICD code for each hospitalization. Therefore, a limitation of our study is that subgroup classification was performed subjectively by the researchers, which may have introduced bias. Additionally, the unavailability of completed VTE protocols prevented verification of whether they were actually filled out by the attending physicians. Thus, it was also not possible to assess whether patients had undergone VTE prophylaxis, either in patients with completed VTE protocols or not.

## CONCLUSIONS

The use of a non-mandatory electronic protocol for managing venous thromboembolism in patients’ electronic records is insufficient to reduce VTE incidence among hospitalized patients across all groups, including Surgical, Clinical, Orthopaedical, and Obstetrical.

## Data Availability

All data produced in the present study are available upon reasonable request to the authors

